# Comparative analysis of cerebrospinal fluid neurofilament medium, light and heavy chain in neurodegenerative diseases using a novel assay for the detection of neurofilament medium chain

**DOI:** 10.1101/2025.01.20.25320650

**Authors:** Badrieh Fazeli, Sara Botzenhardt, Franziska Bachhuber, Paula Klassen, Veronika Klose, Johannes Dorst, Maximilian Wiesenfarth, Zeljko Uzelac, Sarah Jesse, David Brenner, Sarah Anderl-Straub, Albert C. Ludolph, Markus Otto, Jochen Weishaupt, Hayrettin Tumani, Steffen Halbgebauer

**Author notes:** **Corresponding author**: Steffen Halbgebauer, Department of Neurology, Ulm University Hospital, Oberer Eselsberg 45, 89081 Ulm, Germany.

## Abstract

**Objective:** Neurofilaments are key axonal proteins, with neurofilament light (NfL) and heavy (NfH) chain recognized as promising biomarkers for neurodegenerative diseases such as amyotrophic lateral sclerosis (ALS). However, neurofilament medium chain (NfM) remained previously underexplored due to a lack of quantitative assays. In this study, we developed a sensitive immunoassay to measure NfM in cerebrospinal fluid (CSF) and analyzed its levels in ALS, Alzheimer’s disease (AD), frontotemporal dementia (FTD), and Lewy body dementia (LBD). Correlations among neurofilaments and their diagnostic performance were also evaluated.

**Methods:** In this study CSF levels of three neurofilament proteins were measured in 271 participants, including patients with ALS (n=91), AD (n=25), FTD (n=38), LBD (n=18), non-neurodegenerative controls (CTRL, n=51), and 48 individuals initially evaluated for ALS but ultimately diagnosed with other conditions (CTRL.DD).

**Results:** All three neurofilaments were significantly elevated in ALS compared to CTRL and CTRL.DD groups (p<0.0001 for both), with NfM and NfL also increased in FTD (p<0.0001 for both) and AD (NfM, p=0.0017; NfL, p=0.0135 ) compared to CTRL. NfH demonstrated the greatest distinction between ALS and FTD (p<0.0001). Strong correlations were observed among neurofilament subunits, particularly between NfM and NfL (r=0.94, 95% CI: 0.93-0.96, p<0.0001). All neurofilaments effectively distinguished ALS from CTRL and CTRL.DD, with AUC values ranging from 0.92 to 0.99. NfM and NfL showed high accuracy in differentiating AD (NfM, AUC: 0.89; NfL, AUC: 0.90) and FTD (NfM, AUC: 0.91; NfL, AUC: 0.92) from CTRL, while NfH best separated ALS from FTD (AUC: 0.96).

**Conclusion:** This study provides the first quantitative comparison of NfM with NfL and NfH in a neurodegenerative cohort, highlighting its potential diagnostic value. Further research with larger cohorts, longitudinal studies, and investigations into neurofilament distribution in different compartments is needed to clarify the distinct roles of NfM, NfL, and NfH in the diagnosis and treatment of neurological diseases.

## Introduction

Neurofilaments constitute a family of intermediate filament proteins that are essential for the development, structural integrity, and functional maintenance of axons within both the central and peripheral nervous systems. The main neurofilament subunits are the neurofilament heavy (NfH), neurofilament medium (NfM) and the neurofilament light chain (NfL) as well as α-internexin, and peripherin. These subunits interact to form heteropolymers, which collectively contribute to the organization and stability of the neuronal cytoskeleton (1,2). Although these subunits share a conserved tripartite structure, they differ markedly in size and in their specific roles within neurofilament assembly and function. NfL, the smallest (68–70 kDa) and most abundant subunit, forms the core backbone of neurofilaments, providing structural support. Conversely, NfM (145–160 kDa) and NfH (200–220 kDa) feature elongated carboxy-terminal tail domains enriched with phosphorylation sites, which extend as side arms that regulate inter-filament spacing and axonal diameter (3,4).

Neurofilaments, particularly NfL, have emerged as promising biomarkers for a wide range of neurological disorders including neurodegenerative diseases such as amyotrophic lateral sclerosis (ALS) and Alzheimer’s disease (AD). These cytoskeletal proteins, released into cerebrospinal fluid (CSF) and blood upon neuronal damage, serve as sensitive indicators of axonal injury and neurodegeneration (5,6). NfL and phosphorylated NfH (pNfH) are significantly elevated in ALS (7–12), with NfL extensively studied in AD (13–15), frontotemporal dementia (FTD) (16–18), and Lewy body dementia (LBD) (19,20), correlating with cognitive decline and neurodegeneration. While numerous studies have explored NfL and pNfH levels in various neurological diseases, research on NfM levels remains limited as so far, no well validated quantitative assays were previously available. However, non-quantitative protein profiling studies have reported elevated NfM levels in patients with ALS and FTD (21–26).

In this study we developed a novel quantitative and highly sensitive immunoassay to measure NfM levels in CSF and subsequently utilize it for the analysis of NfM in the CSF of patients with neurological disorders, including ALS, AD, FTD, and LBD, alongside control cohorts. Additionally, we investigated correlations between the NfM, NfL and NfH and employed receiver operating characteristic (ROC) analysis to evaluate and compare their individual discriminating potentials.

## Method

### Patient selection

The focus of this study was to measure the levels of three neurofilament proteins in the CSF of 271 patients diagnosed with ALS (n=91), AD (n=25), FTD (n=38), LBD (n=18), and two control groups consisting of (i) individuals initially under suspicion of ALS but finally diagnosed differently (CTRL.DD) (n=48) (see Table S1 for diagnoses) and (ii) non-neurodegenerative controls (CTRL) (n=51).

ALS Patients met the criteria for definite or probable ALS based on the revised El Escorial criteria (27). The diagnosis of AD in patients was established based on the International Working Group 2 criteria (28). This diagnosis was further supported by the analysis of core CSF biomarkers (A: Abeta 42 to 40 ratio, T: phosphorylated tau 181 (ptau 181), N: total tau), following the recommendations from the National Institute on Aging and Alzheimer’s Association (29). All AD cases presented a CSF biomarker profile of A+T+N+. The FTD group included 17 patients diagnosed with behavioral variant frontotemporal dementia (bvFTD), along with 21 patients with primary progressive aphasia (PPA) subtypes: 7 with the non-fluent variant, 7 with the logopenic variant, and 7 with the semantic variant. Diagnoses of bvFTD and PPA were made in accordance with accepted international criteria (30,31).

Among the 18 patients with LBD, 14 were diagnosed with Parkinson’s disease (PD) based on the UK Parkinson’s Disease Society Brain Bank criteria (32), while 4 were diagnosed with Parkinson’s disease dementia (PDD) based on significant impairment in daily functioning (33) and the clinical criteria for PDD recommended by the Movement Disorder Society (34).

The CTRL.DD group comprised patients initially suspected of ALS but later diagnosed with other conditions, with their NfL and NfH levels previously assessed by Halbgebauer et al (12).

The CTRL group included control patients with no clinical signs of neurodegeneration. These subjects were initially admitted to the department of neurology due to symptoms such as tension-type headaches, brief sensory disturbances or dizziness. Thorough clinical and radiological examinations ruled out neurodegenerative and neuroinflammatory disorders. Each control participant also underwent a lumbar puncture to rule out possible central nervous system (CNS) inflammation. Evaluation criteria included a normal leukocyte count, preserved blood-CSF barrier function (reflected by a normal CSF albumin-to-serum ratio) and no evidence of intrathecal immunoglobulin synthesis, confirmed by quantitative analysis of IgG, IgA, IgM and oligoclonal IgG bands.

CSF samples were collected at the Department of Neurology, University Hospital of Ulm, Germany, between 2010 and 2021. All participants or their legal representatives gave informed consent to participate in the study. The study was approved by the Ethics Committee of the University of Ulm (approval number: 20/10) and was conducted in accordance with the Declaration of Helsinki.

### CSF sampling and analysis

CSF samples were obtained by lumbar puncture, centrifuged at 2,000g for 10 minutes, and the supernatant was aliquoted and stored at -80°C (35). To quantify NfL and NfH in CSF samples, commercially available Ella microfluidic kits (Bio-techne, Minneapolis, USA) were used and measurements were performed according to the manufacturer’s instructions. The quantification range for these assays is 2.7 to 10,290 pg/mL for NfL and 7.46 to 28,480 pg/mL for NfH. CSF NfM levels were measured using a newly developed sandwich ELISA assay.

### Antibodies and recombinant protein

The novel NfM immunoassay included a mouse monoclonal antibody, clone OTI2C4 (Cat. # CF506794, OriGene Technologies, Rockville, MD, USA) against NfM as capture and a mouse monoclonal antibody, clone OTI2G3 (Cat. # NBP2-72977, Novus Biologicals, Littleton, Colorado, USA) as detector. The detector antibody was biotinylated with EZ-Link™ NHS-PEG4-Biotin (Cat. # A39259, Thermo Fisher Scientific, Massachusetts, USA), in a ratio biotin to antibody 40:1 according to the biotinylation protocol provided by Quanterix Corporation (Lexington, Massachusetts, USA). For assay development and antibody affinity screening, human recombinant NfM (Cat. # TP324475, OriGene Technologies, Rockville, MD, USA) was employed.

### NfM Sandwich ELISA method

Nunc Maxisorp 96-well microtiter plates (Thermo Fisher Scientific, Massachusetts, USA) were coated with 100 µL per well of capture antibody (Cat. # CF506794) at a concentration of 3.3 µg/mL in 100 mM bicarbonate-carbonate buffer (pH 9.6) and incubated overnight at 4°C. Following removal of the coating solution, non-specific binding sites were blocked by adding 300 µL of blocking buffer (1% bovine serum albumin in phosphate-buffered saline (PBS) with 0.05% Tween 20) to each well, followed by incubation at 20°C for 2 hours. CSF samples were diluted 1:4 in blocking buffer, and calibrators were prepared using recombinant NfM (Cat. #TP324475) with concentrations ranging from 125 to 8000 pg/mL. A volume of 100 µL of the diluted CSF samples, blocking buffer as blank, controls and calibrators was added in duplicate and incubated at 30°C for 1.5 hours. The wells were then washed three times with 300 µL of wash buffer (PBS with 0.05% Tween 20) to remove unbound proteins. Subsequently, 100 µL of biotinylated detector antibody (Cat. #NBP2-72977), diluted to 1.32 µg/mL in blocking buffer, was applied to each well and incubated for 1 hour at 20°C. After further washing, 100 µL of avidin/biotin-based peroxidase complexes (A& B solutions,1:200 each in PBS) (Cat. #PK-6100, Vector Laboratories, California, USA) was added and incubated for 1 hour at 20°C to allow detection. The plate was washed again and 100 µL of 3,3′,5,5′-tetramethylbenzidine (Thermo Fisher Scientific, Massachusetts, USA) was added to each well and incubated for 15 minutes at room temperature in the dark to allow color development. The reaction was stopped by adding 100 µL of 1M hydrochloric acid to each well. Absorbance was measured at 450 nm with a reference wavelength of 570 nm. Concentrations were determined from a 4-parameter logistic standard curve.

### Novel NfM assay validation

Repeatability was evaluated by measuring eight replicates of a pooled CSF sample along with two individual CSF samples. To assess intermediate precision, four replicates of three individual CSF samples and one pooled sample were measured across three separate runs. The lower limit of quantification (LLOQ) and the limit of detection (LOD) were determined using 16 blank measurements, with the LLOQ calculated as the signal corresponding to 10 standard deviations (SD) above the mean, and the LOD calculated as 3 SD above the mean (36).

The calibrators covered a concentration range of 125 to 8,000 pg/mL. 5% of the samples measured exceeded the upper limit and their concentration was estimated by extrapolation. All samples were above the LLOQ. To assess parallelism, five endogenous CSF samples were analyzed - one with a high concentration and four with low concentrations - diluted at ratios ranging from 1:2 to 1:8. The back-calculated concentrations from these dilutions were evaluated to determine the minimum required dilution (MRD). This strategy was implemented to minimize matrix effects and ensure accurate quantification of endogenous NfM.

To assess spike and recovery, two CSF samples were diluted at a ratio of 1:4 and divided into three aliquots. Each aliquot was then spiked with NfM-free sample diluent, as well as recombinant NfM protein (Cat. # TP324475) at medium (2,000 pg/mL) and low (400 pg/mL) concentrations. The volume of the spiked solution was kept below 10% of the total aliquot volume, and recovery was expressed as a percentage. To evaluate potential cross-reactivity with abundant CSF proteins, serial dilutions of the two CSF samples were spiked with physiological concentrations of human serum albumin (HSA) (200 µg/mL) and a higher concentration of 600 µg/mL, as well as physiological concentrations of immunoglobulin G (IgG) (30 µg/mL) and a higher concentration of 90 µg/mL. NfM levels in these spiked samples were then compared to those in unspiked samples.

To assess potential cross-reactivity with NfL and NfH in an indirect ELISA, recombinant proteins for NfL (Cat. # ab224840, Abcam, Cambridge, UK) and NfH (Cat. # TP313487, OriGene Technologies, Rockville, MD, USA) were coated onto the assay plate, and antibodies were screened for their affinity to these two proteins. Antibodies specific to NfL (Cat. # 130400, Thermo Fisher Scientific, Massachusetts, USA) and NfH (Cat. # 18934-1-AP, Proteintech, Rosemont, IL, USA) were used as positive controls. Furthermore, NfL and NfH recombinant proteins were used as samples in the newly developed sandwich ELISA to further evaluate possible cross-reactivity.

### Statistical methods

Data analysis and visualization were performed using GraphPad Prism software, version 10.2.2 (GraphPad Software, La Jolla, CA, USA). The Shapiro–Wilk test was conducted to assess the data distribution. Since the data did not follow a Gaussian distribution, non-parametric tests were applied. Neurofilament concentrations were normalized using z-scores. To calculate the Z-scores, the absolute values were first log2-transformed, and then the following formula was applied: Z = (X − μ_controls) / σ_controls. In this equation, X represents each individual value within the patient cohort, μ_controls is the mean value of the control group, and σ_controls is the standard deviation of the control group. Mann-Whitney U tests were used to assess significant differences between groups for pairwise comparisons, and the Kruskal-Wallis test followed by Dunn’s post-hoc analysis was used for comparisons between multiple groups. Spearman correlation coefficients were calculated to assess the correlations among neurofilament proteins and their association with age. ROC analyses were performed to determine cut-off values. Statistical significance was defined as p < 0.05.

## Results

### Performance of the established ELISA assay for the detection of NfM

The newly developed assay, targeting full-length recombinant NfM protein, demonstrated intra- and inter-assay variability of 5.5% and 10%, respectively. The LLOQ and LOD of the assay were determined to be 107.7 pg/mL and 23.9 pg/mL, respectively, with no cross-reactivity observed with NfL or NfH (Figure S1). Parallelism experiments indicated a minimum required dilution (MRD) of 1:2; however, CSF samples were diluted 1:4 in subsequent measurements to maximize the number of samples within the range of the calibration curve (Figure S2). Recovery analysis of the recombinant protein spiked at a 1:4 dilution showed a recovery rate of 96%. Stability testing showed that CSF NfM is stable for up to three days at room temperature or 4°C, and up to five freeze-thaw cycles did not affect measured NfM concentrations, with variability remaining below 20%. No cross-reactivity with human albumin or immunoglobulin G was detected. Further details of assay performance are provided in the Supplementary Materials.

### Demographic features and neurofilament protein concentrations

The main demographic parameters for each diagnostic group are summarized in Table 1. No significant difference in age was observed between the groups. There was also no significant difference between neurofilament levels in female and male control patients (NfM (p=0.83), NfL (p=0.83), NfH (p=0.5)). Correlation analysis revealed a strong correlation between NfM concentrations and age in CTRL (r=0.69 (95% CI: 0.51 to 0.81), p<0.0001), and CTRL.DD (r=0.77 (95% CI: 0.62 to 0.87), p<0. 0001), but not in the patients cohorts (Figure 1).

**Figure 1.**
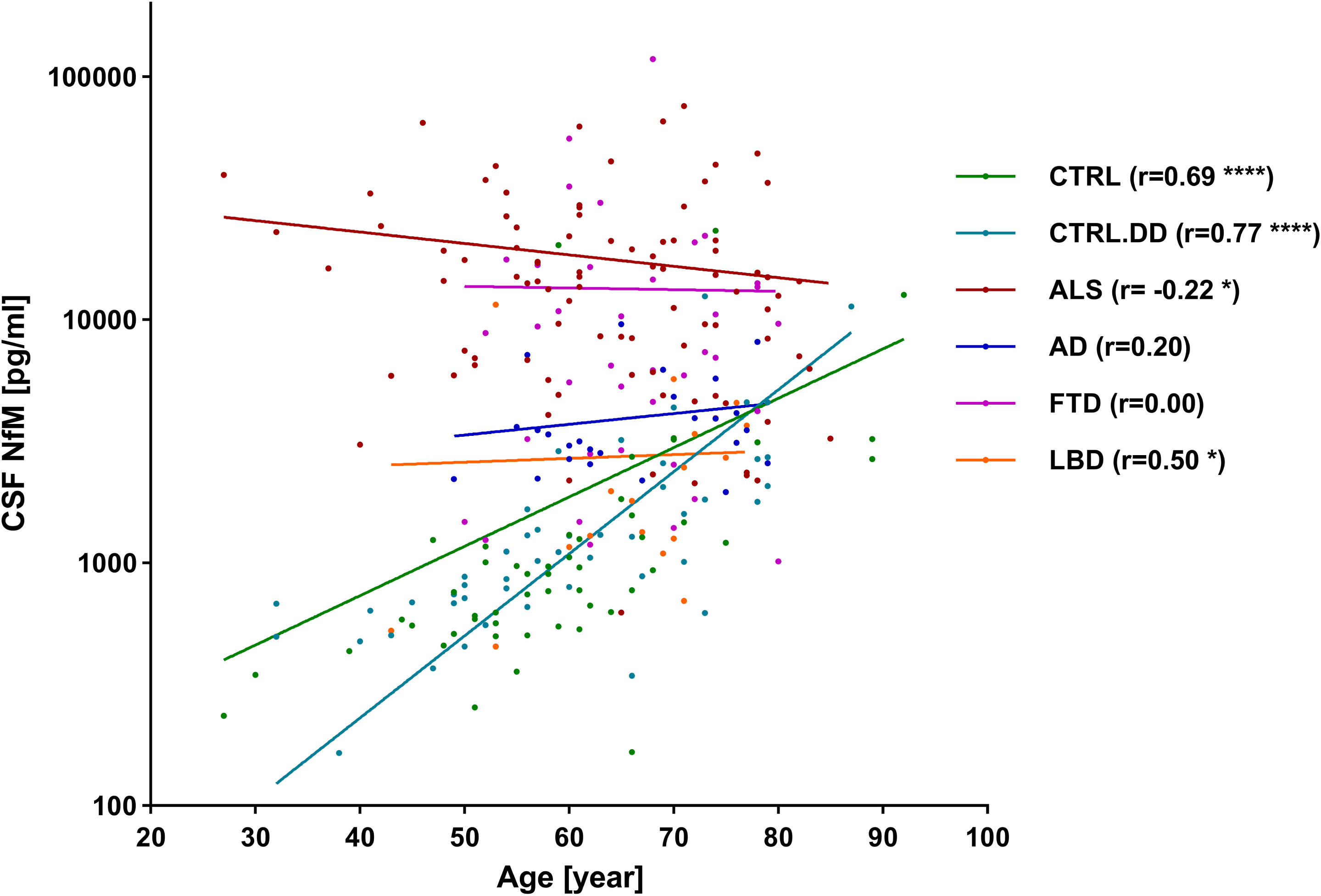
Assessment of correlation between CSF NfM and age. Spearman correlation between CSF NfM and age in each diagnostic cohort is shown in different colors. CTRL (r=0.69 (95% CI: 0.51 to 0.81), p<0.0001), CTRL.DD (r=0.77 (95% CI: 0.62 to 0.87), p<0. 0001), ALS (r= -0.22 (95% CI: -0.42 to -0.01), p=0.03), AD (r=0.20 (95% CI: -0.22 to 0.56), p=0.33), FTD (r=0.00 (95% CI: -0.32 to 0.33), p= 0.97), and LBD (r=0.50 (95% CI: 0.03 to 0.79), p=0.03). AD, Alzheimer’s disease; ALS, amyotrophic lateral sclerosis; CTRL, non-neurodegenerative controls; CTRL.DD, control patients with initial diagnostic suspicion of ALS but finally diagnosed with another condition; CSF, cerebrospinal fluid; FTD, frontotemporal dementia; LBD, lewy body dementia; NfM, neurofilament medium chain.

**Table 1.** Demographic data of the diagnostic cohort.

|  | Control |  | ALS | AD | FTD |  |  |  | LBD |  |
| --- | --- | --- | --- | --- | --- | --- | --- | --- | --- | --- |
| Subgroups | CTRL | CTRL.DD | - | - | bvFTD | lvPPA | nfvPPA | svPPA | PD | PDD |
| N | 51 | 48 | 91 | 25 | 17 | 7 | 7 | 7 | 14 | 4 |
| female/male | 26/25 | 15/33 | 44/47 | 17/8 | 4/13 | 3/4 | 3/4 | 2/5 | 5/7 | 1/3 |
| Age at LP [year] | 59 (52-66) | 59 (50-70) | 64 (55-74) | 65 (59-74) | 63 (60-69) | 71(62-73) | 73(60-78) | 65(57-74) | 66(60-71) | 75(71-77) |
| CSF NfM[pg/mL] | 900 (552-1304) | 1034 (678-1989) | 14425 (6512-22964) | 3377 (2619-4478) | 5308(1432-18613) | 4590(2806-6972) | 14106(9380-22132) | 10854 (10311-16793) | 1883(1194-3793) | 3188 (1620-4337) |
| CSF NfL[pg/mL] | 563 (449-779) | 592 (446-890) | 6033 (4092-9207) | 1344 (1064-1650) | 1528(1006-3154) | 1528(1324-2208) | 3528(2440-5788) | 3272(2840-5032) | 966(589-1488) | 1626(1379-1732) |
| CSF NfH[pg/mL] | 756 (412-1328) | 953 (712-1302) | 6324 (4137-9476) | 1112 (702-1376) | 1084 (726-2076) | 1500(1064-1892) | 1904(1208-2236) | 808(496-1424) | 1202(816-1868) | 2024(1415-2270) |

Data is reported as median (Interquartile range).
Abbreviations: AD, Alzheimer's disease; ALS, amyotrophic lateral sclerosis; bvFTD, behavioral variant frontotemporal dementia; CTRL, non-neurodegenerative controls; CTRL.DD, control patients with initial diagnostic suspicion of ALS but finally diagnosed with another condition; CSF, cerebrospinal fluid; FTD, frontotemporal dementia; LBD, lewy body dementia; lvPPA, logopenic variant primary progressive aphasia; NfH, neurofilament heavy chain; NfL, neurofilament light chain; NfM, neurofilament medium chain; nfvPPA, non-
fluent variant primary progressive aphasia; PD, Parkinson's disease; PDD, Parkinson's disease dementia; svPPA, semantic variant primary progressive aphasia.

### NfM, NfL and NfH levels in the diagnostic groups

All three neurofilaments showed significantly elevated levels in ALS compared with CTRL and CTRL.DD with p<0.0001 (Figure 2A). However, NfM and NfL were significantly increased in FTD vs CTRL (p<0.0001 for both) and AD vs CTRL (NfM, p=0.0017; NfL, p=0.0135 ), whereas NfH values revealed no significant difference. NfH on the other hand displayed the highest difference between ALS and FTD (p<0.0001). To further assess differences in neurofilament protein concentrations, we compared the levels across disease subgroups (Figure 2B).

**Figure 2.**
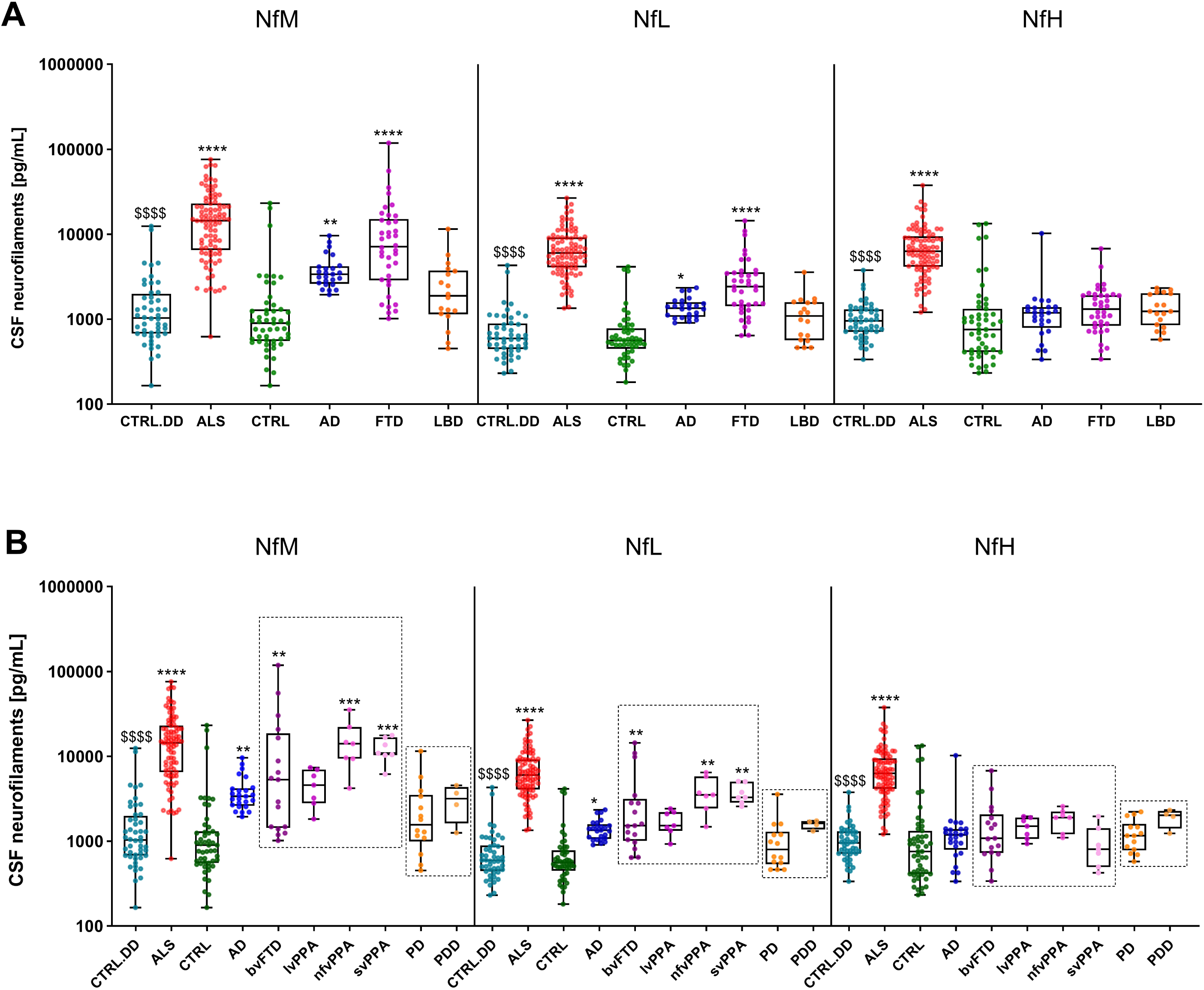
Neurofilament proteins in diagnostic groups. CSF NfM, NfL and NfH concentrations in the diagnostic groups (A) and in the extended diagnostic groups (B). Statistically significant differences between the patient cohorts and CTRL are indicated with star symbols (*), while the comparison between ALS patients and the CTRL.DD cohort is marked with dollar signs ($). Additional significant differences within the patient cohorts are observed with the following p-values; NfM (ALS vs AD: p=0.0017; ALS vs LBD: p<0.0001; FTD vs LBD: p=0.0179), NfL (ALS vs AD: p<0.0001; ALS vs FTD: p= 0.0016; ALS vs LBD: p<0.0001), and NfH (ALS vs AD: p<0.0001; ALS vs FTD: p<0.0001; ALS vs LBD: p<0.0001). Displayed are the median concentration, the 25% and 75% percentiles and whiskers from minimum to maximum. Groups were compared by Kruskal-Wallis test and Dunns post hoc test.( * p< 0.05; ** p< 0.01; *** p< 0.001; **** p<0.0001; and $$$$ p<0.0001). AD, Alzheimer’s disease; ALS, amyotrophic lateral sclerosis; bvFTD, behavioral variant frontotemporal dementia; CTRL, non-neurodegenerative controls; CTRL.DD, control patients with initial diagnostic suspicion of ALS but finally diagnosed with another condition; CSF, cerebrospinal fluid; FTD, frontotemporal dementia; LBD, lewy body dementia; lvPPA, logopenic variant primary progressive aphasia; NfH, neurofilament heavy chain; NfL, neurofilament light chain; NfM, neurofilament medium chain; nfvPPA, non-fluent variant primary progressive aphasia; PD, Parkinson’s disease; PDD, Parkinson’s disease dementia; svPPA, semantic variant primary progressive aphasia.

A pairwise comparison was made within the FTD and LBD subgroups. Significantly lower NfM levels were observed in lvPPA compared to nfvPPA (p = 0.007) and svPPA (p = 0.0023). Similarly, NfL levels were significantly lower in lvPPA compared to nfvPPA (p = 0.007) and svPPA (p = 0.0006). However, no significant differences were found for NfH levels. All three proteins displayed a trend to elevated levels in the PDD group compared to PD, with only NfL levels showing a significant difference (p = 0.034). Although the absolute values of the three neurofilament proteins are presented in the same graph (Figure 2), direct comparison is not possible due to the use of different assays, each calibrated independently. To enable comparison of variations among the proteins, the absolute values were log transformed, and z-scores were calculated. These normalized data are displayed in Figure 3. Additionally, we calculated ratios between different neurofilament proteins and compared these ratios across diagnostic groups; detailed results of these analyses are provided in the supplementary material (Figure S3).

**Figure 3.**
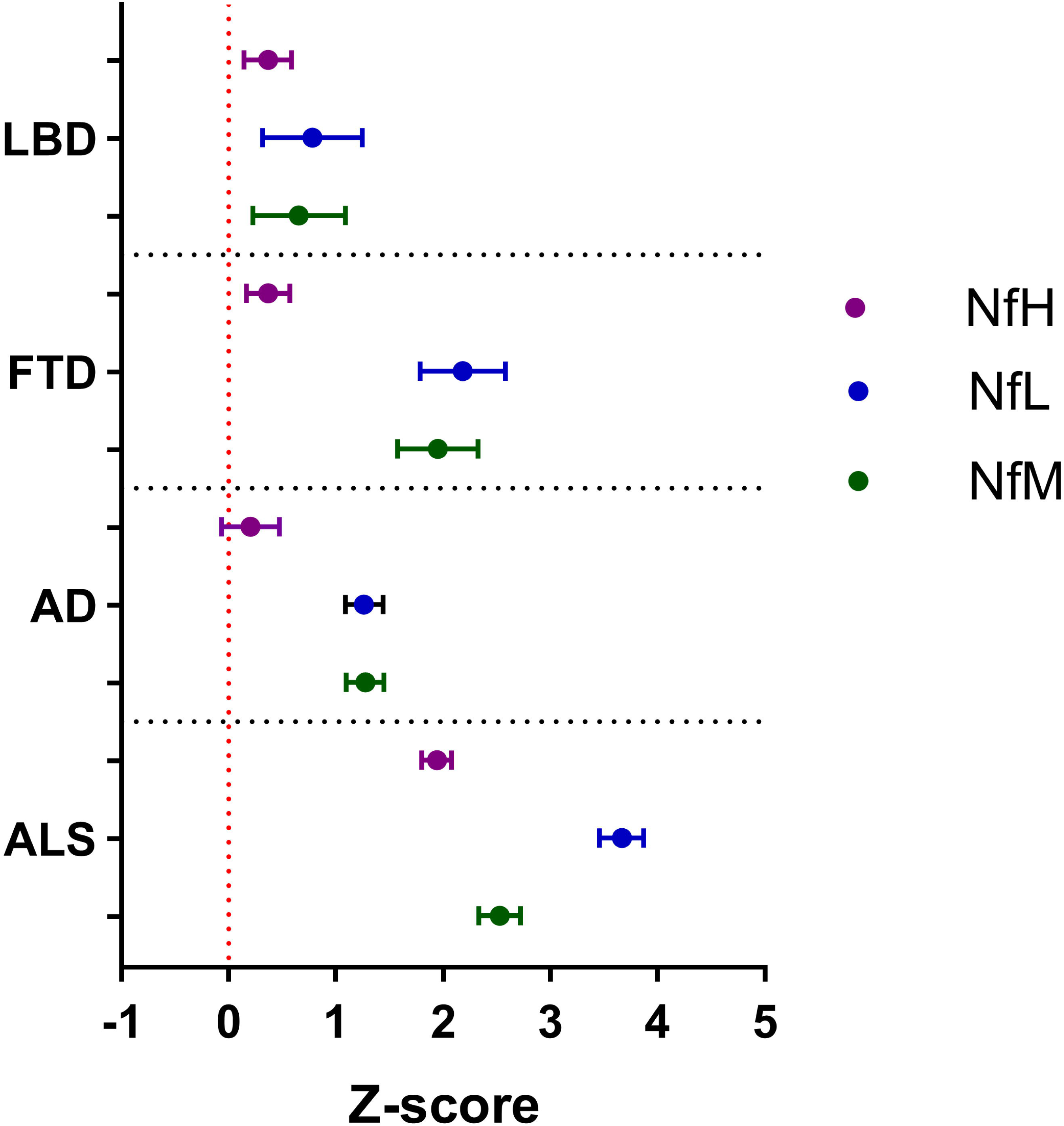
Neurofilament protein comparison using z-scores. NfL, NfM and NfH levels were normalized using z-scores and visualized in a forest plot layout. The forest plot displays the mean z-scores and corresponding 95% confidence intervals, illustrating the variations in different proteins values within each patient cohort compared to the mean value in the respective control cohort. The NfM values are depicted in green, NfL in blue, and NfH in purple. AD, Alzheimer’s disease; ALS, amyotrophic lateral sclerosis; FTD, frontotemporal dementia; LBD, lewy body dementia; NfH, neurofilament heavy chain; NfL, neurofilament light chain; NfM, neurofilament medium chain.

### Associations of CSF neurofilaments with each other and ATN scores

In the entire cohort, CSF NfM values showed a stronger correlation with CSF NfL (r=0.94, 95% CI: 0.93-0.96, p<0.0001) (Figure 4A), than with CSF NfH (r=0.72, 95% CI: 0.66-0.78, p<0.0001) (Figure 4B). A moderate to strong correlation was noted between CSF NfL and CSF NfH (r=0.79, 95% CI: 0.74-0.83, p<0.0001) (Figure 4C). Analysis of ATN scores and neurofilament proteins revealed no significant correlation between the amyloid-beta ratio and any protein (Figure 5A). However, NfM showed a strong and significant correlation with both pTau181(r=0.70 (95% CI: 0.40 to 0.86), p=0.0001) (Figure 5B), and total tau (r=0.80 (95% CI: 0.58 to 0.91), p<0.0001) (Figure 5C).

**Figure 4.**
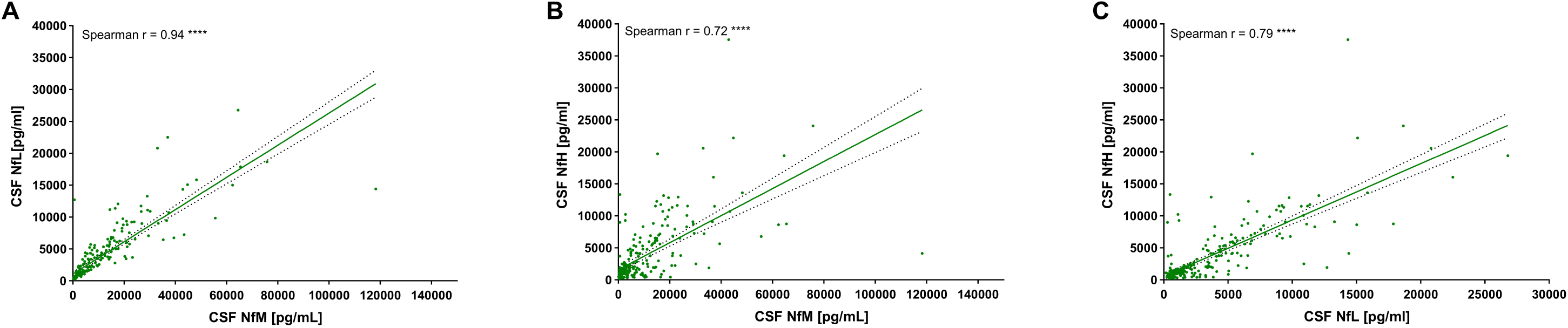
Correlation analysis between the different neurofilament proteins. (A) Correlation between CSF NfM and NfL (r=0.94 (95% CI 0.93 to 0.96), p<0.0001) (n=271). (B) Correlation between CSF NfM and NfH (r=0.72 (95% CI 0.66 to 0.78), p<0.0001) (n=271). (C) Correlation between CSF NfL and NfH (r=0.79 (95% CI 0.74 to 0.83), p<0.0001) (n=271). Correlation analysis was performed using Spearman’s correlation coefficient. CI, confidence intervals; CSF, cerebrospinal fluid; NfH, neurofilament heavy chain; NfL, neurofilament light chain; NfM, neurofilament medium chain.

**Figure 5.**
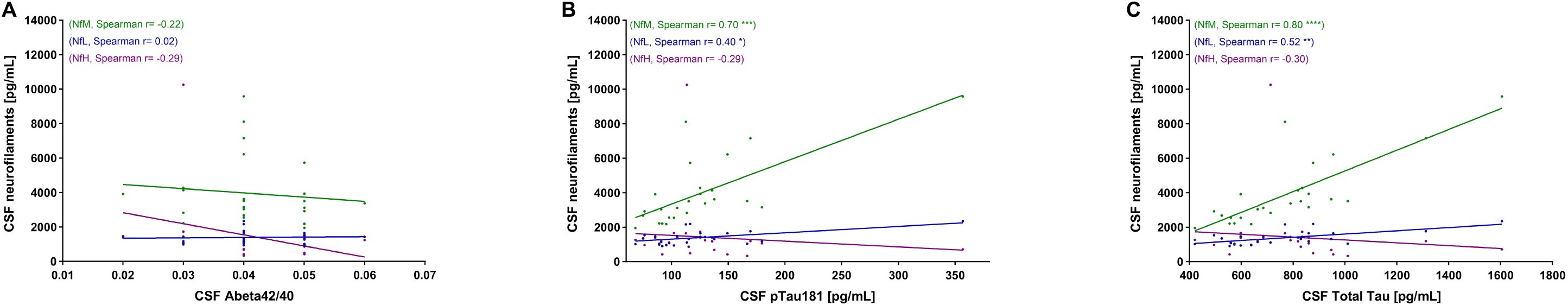
Correlation analysis between ATN scores and CSF neurofilaments in AD cohort. (A) Correlation between Amyloid beta 42/40 and NfM (r= -0.22 (95% CI -0.57 to 0.20), p=0.29), n=25), NfL (r= 0.02 (95% CI -0.39 to 0.42), p=0.93), n=25), NfH (r= -0.29 (95% CI -0.62 to 0.13), p=0.16), n=25) (B) Correlation between pTau 181 and NfM (r= 0.70 (95% CI 0.40 to 0.86), p=0.0001), n=25), NfL (r= 0.40 (95% CI -0.01 to 0.70), p=0.0492), n=25), NfH (r= -0.29 (95% CI -0.62 to 0.13), p=0.1562), n=25) (C) Correlation between Total Tau and NfM (r= 0.80 (95% CI 0.58 to 0.91), p<0.0001), n=25), NfL (r= 0.52 (95% CI 0.14 to 0.76), p=0.0081), n=25), NfH (r= -0.30 (95% CI -0.63 to 0.11), p=0.1380), n=25). Correlation analysis was performed using Spearman’s correlation coefficient. CI, confidence intervals; CSF, cerebrospinal fluid; NfH, neurofilament heavy chain; NfL, neurofilament light chain; NfM, neurofilament medium chain.

### Discriminative potential of CSF neurofilament proteins

All three neurofilament proteins could effectively discriminate ALS from the CTRL patients with an area under the curve (AUC) of 0.95 for NfM, 0.98 for NfL and 0.92 for NfH (Figure 6A). Similarly, ALS patients could be well distinguished from the CTRL.DD cases by NfM (AUC: 0.96), NfL (AUC: 0.99) and NfH (AUC: 0.99) (Figure 6B). In addition, NfM and NfL revealed higher accuracy for the discrimination between AD (NfM, AUC: 0.89; NfL, AUC: 0.90) (Figure 6C) and FTD (NfM, AUC: 0.91; NfL, AUC: 0.92) (Figure 6D) patients and CTRL cases. For the discrimination between ALS and FTD patients, NfH showed the best result (AUC: 0.96), followed by NfL (AUC: 0.83) (Figure 6E). However, the performance of neurofilament proteins was suboptimal for discriminating between AD and FTD patients (Figure 6F). The combination of neurofilament proteins did not enhance the accuracy of discrimination between patient cohorts (data not shown).

**Figure 6.**
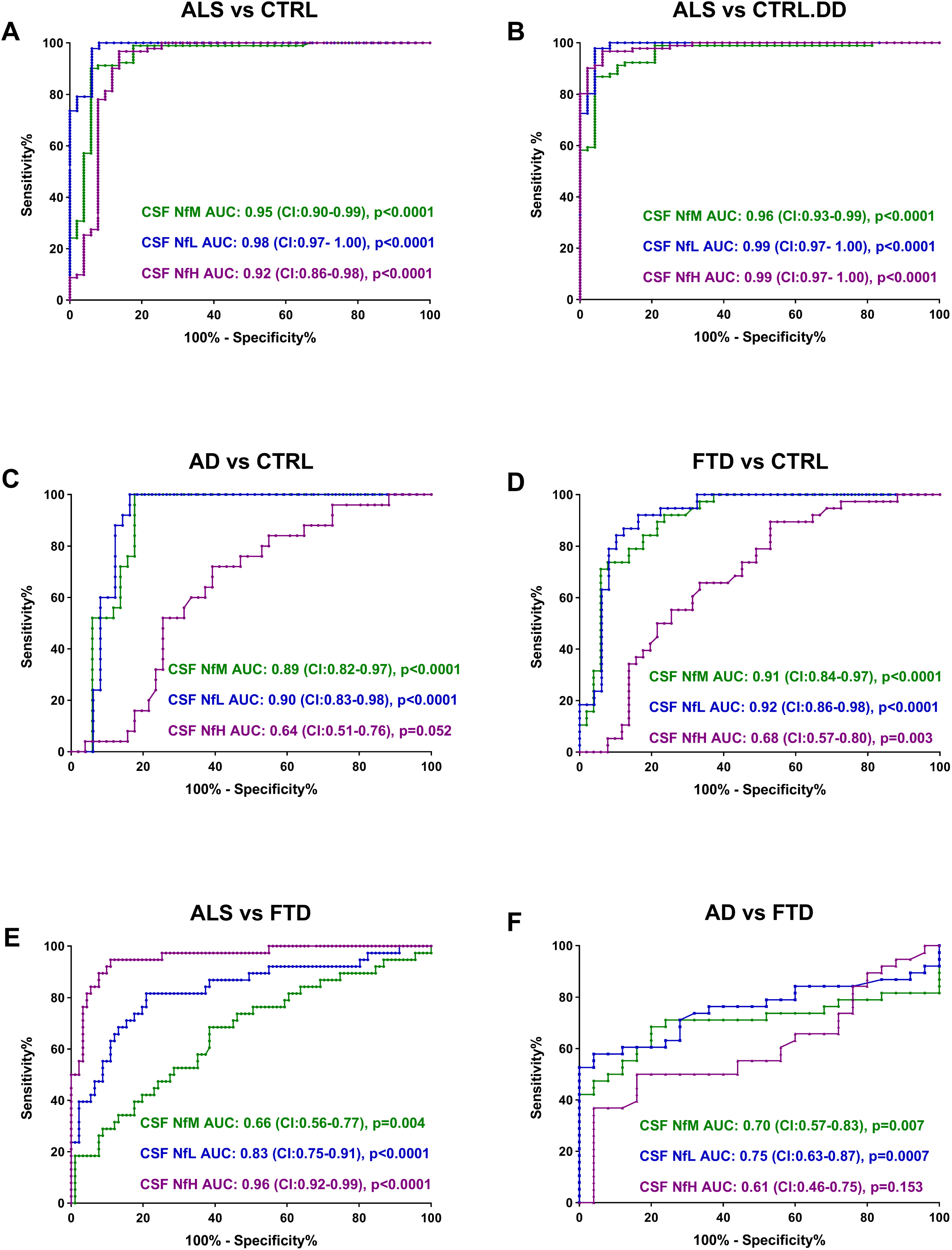
ROC analysis of CSF neurofilament proteins. The panels show the results of the ROC analyses comparing the levels of three neurofilament proteins in the CSF of the following groups: (A) ALS vs. CTRL, (B) ALS vs. CTRL.DD, (C) AD vs. CTRL, (D) FTD vs. CTRL, (E) ALS vs. FTD, and (F) AD vs. FTD. AD, Alzheimer’s disease; ALS, amyotrophic lateral sclerosis; CI, confidence intervals; CTRL, non-neurodegenerative controls; CTRL.DD, control patients with initial diagnostic suspicion of ALS but finally diagnosed with another condition; CSF, cerebrospinal fluid; FTD, frontotemporal dementia; NfH, neurofilament heavy chain; NfL, neurofilament light chain; NfM, neurofilament medium chain; NfL, neurofilament light chain protein; ROC, receiver operating characteristic.

## Discussion

CSF NfL and NfH are well studied in the literature as axonal damage markers. However, the third neurofilament NfM is lagging behind in terms of available and well validated assays and analyses in neurological diseases. In this study we developed a novel sensitive ELISA for the quantification of NfM in CSF and applied it in a comprehensive cohort of neurodegenerative diseases. The technical validation demonstrated an excellent assay performance meeting all relevant technical criteria. Of particular note is the high specificity for NfM with no cross-reactivity to NfL and NfH.

Our study is the first one to use a well validated quantitative assay for NfM and compare CSF levels between NfM, NfL and NfH. As both NfL and NfH correlate strongly with age (6,37,38) we first examined if NfM (and also NfL and NfH) depicts the same association. Confirming the literature NfL and NfH illustrated a strong positive association with age in the control group which we also detected for NfM. This effect has to be taken into account when analyzing patients in different age groups. In our cohort of control and neurodegenerative diseases there was no significant difference in age. NfM, NfL and NfH on the other hand showed significantly elevated concentrations in ALS compared to control and CTRL.DD patients. While elevated levels of NfL and NfH in ALS have been well-documented (10,12,39), quantitative data for NfM has been lacking. Our study supports recent findings demonstrating elevated NfM levels in ALS using a semi-quantitative bead suspension array (40).

Furthermore, we could demonstrate a significant elevation of NfM and NfL but not NfH in AD and FTD compared to control patients which was in the case of AD most significant for NfM. This finding confirms the literature showing better discriminating potential for NfL than NfH for AD and FTD compared to controls (41–44). The NfM results now complement these findings. On the other hand, only NfH was elevated in ALS compared to FTD patients corroborating findings of other studies (12,17,45,46). In the FTD subgroups, we observed significantly elevated NfM and NfL levels in patients with nfv and svPPA compared to those with bvFTD and lvPPA, a finding previously reported only for NfL (47).

Taken together, NfM levels in CSF are more comparable to NfL CSF concentrations than to NfH, despite NfM being more closely related to NfH in terms of amino acid sequence, structure and neurofilament assembly (4). This observation is supported by the correlation analysis between the three neurofilaments, which revealed the strongest association between NfL and NfM. In contrast, NfL and NfH showed a moderate to strong association, consistent with findings from previous studies (12,48).

Given the prominently elevated NfM levels in the AD cohort, we also analyzed its correlation with the CSF ATN biomarkers assessed in these patients. Notably, NfM showed the strongest association with p-tau181 and t-tau. While the AD group included only 20 patients, limiting definitive conclusions, this finding warrants further investigation of NfM in a larger AD cohort.

NfM showed similar results to NfL and NfH in discriminating between disease and controls. We confirmed recent semi-quantitative analyses showing high AUCs for NfM in discriminating ALS patients from control patients (49), findings are also well-documented for NfL and NfH (7,8,46,50). NfM also exhibited strong discriminatory potential in Alzheimer’s disease patients, similar to NfL, in differentiating AD from controls (51–53). However, NfM did not enhance the diagnostic power of neurofilaments in the neurodegenerative groups tested. The strength of our study lies in the use of the first well characterized and validated quantitative immunoassay for detection of NfM in CSF. Furthermore, we evaluated a comprehensive cohort of neurodegenerative diseases, with parallel assessment of NfL and NfH for comparison. However, limitations include the cross-sectional design, which did not allow us to track NfM changes over time, and the relatively small sample sizes in some subgroups.

In conclusion, we present the first quantitative comparison of NfM with NfL and NfH in a neurodegenerative cohort, contributing to the existing body of literature on NfL and NfH. Further studies on NfM, especially in FTD and AD with larger patient cohorts, as well as investigations of NfM in neuroinflammatory diseases and in longitudinal studies will provide more insight into its potential value for the (differential-) diagnosis and monitoring of neurological diseases. Additionally, given the apparent differences in the CSF concentration patterns of NfM, NfL and NfH across neurological diseases, studies examining the expression and distribution of these neurofilaments in different brain compartments could help clarify their distinct roles in various neurological disorders.

## Data Availability

All data produced in the present study are available upon reasonable request to the corresponding author.

## Acknowledgments

We extend our gratitude to all the patients who participated in this study, to their caregivers, and to the healthcare professionals involved in the care of patients. We also thank the biobank of the Department of Neurology in Ulm (Alice Beer, Sandra Hübsch and Dagmar Schattauer) for their help with providing the samples.

## Competing Interests

HT reports honoraria for acting as a consultant/speaker and/or for attending events sponsored by Alexion, Bayer, Biogen, Bristol-Myers Squibb, Celgene, Diamed, Fresenius, Fujirebio, GlaxoSmithKline, Horizon, Janssen-Cilag, Merck, Novartis, Roche, Sanofi-Genzyme, Siemens, Teva and Viatris. All conflicts are not relevant to the topic of the study. All other authors declare no competing interests.

## Study Funding

The present study was supported by the Else Kröner Fresenius Foundation and Chemische Fabrik Karl Bucher GmbH.

## Author Contributions

All authors made significant contributions to the conception and design of the study, and/or the acquisition, analysis, and interpretation of data. Each author has reviewed and approved the final version of the manuscript for submission and accepts responsibility for all aspects of the work, ensuring that any concerns regarding the accuracy or integrity of any part of the study are thoroughly investigated and appropriately addressed.

Conception and design of the study: BF, HT, SH; Sample collection and data management: BF, SB, FB, PK, VK, JD, MW, ZU, SJ, DB, SAS, ACL, MO, JW, HT, SH; Study management and coordination: BF, FB, HT, SH; Statistical methods and analysis: BF, HT, SH; Interpretation of results: BF, MO, HT, SH; Manuscript writing (first draft): BF, SH; Critical revision of the manuscript: BF, SB, FB, PK, VK, JD, MW, ZU, SJ, DB, SAS, ACL, MO, JW, HT, SH.

## Notes

### Funding Statement

The present study was supported by the Else Kroener Fresenius Foundation and Chemische Fabrik Karl Bucher GmbH.

### Author Declarations

The study was approved by the Ethics Committee of the University of Ulm (approval number: 20/10) and was conducted in accordance with the Declaration of Helsinki.

