## Supplementary material for "Comparative analysis of cerebrospinal fluid neurofilament medium, light and heavy chain in neurodegenerative diseases using a novel assay for the detection of neurofilament medium chain"

**Corresponding author**

### Supplementary material

#### Cross reaction with NfL and NfH

Given the structural similarities between the neurofilament subunits, we investigated the potential for cross-reactivity between NfL, NfM and NfH. To this end, we tested the affinity of all antibodies for recombinant NfM (Cat. # TP324475), NfL (Cat. # ab224840) and NfH (Cat. # TP313487) using an indirect ELISA where the recombinant protein was coated to the bottom of the wells (data are not shown). Results are presented for the two antibodies used in the final assay (capture antibody, Cat. # CF506794; detector antibody, Cat. #NBP2-72977). To confirm the presence of recombinant NfL and NfH, specific NfL and NfH antibodies were included in the measurements (NfL antibody, Cat. # 130400; NfH antibody, Cat. # 18934-1-AP).

In the indirect ELISA the capture and detector NfM antibodies generated strong signals for NfM and signals for NfL and NfH at the blank level. Only the capture antibody also showed a bottom-line signal for NfH in the indirect ELISA. However, when we assessed the cross-reactivity of the final antibody combination by using all three recombinant proteins as samples in the developed sandwich ELISA, zero cross reaction with NfL and NfH was detected (Figure S1).

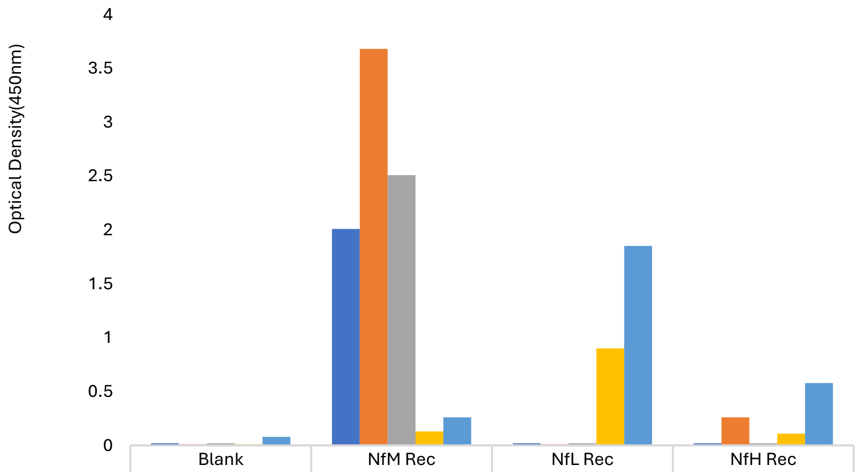


| | Blank | NfM Rec | NfL Rec | NfH Rec |
|---|---|---|---|---|
| Sandwich ELISA- Final assay | 0.02 | 2.01 | 0.02 | 0.02 |
| Indirect ELISA- Capture Ab (CF506794) | 0.01 | 3.68 | 0.01 | 0.26 |
| Indirect ELISA- Detector Ab (NBP2-72977) | 0.02 | 2.51 | 0.02 | 0.02 |
| Indirect ELISA- NfL Ab (13-0400) | 0.01 | 0.13 | 0.9 | 0.11 |
| Indirect ELISA- NfH Ab (18934-1-AP) | 0.08 | 0.26 | 1.85 | 0.58 |

**Figure S1. Assessment of cross reactivity with other neurofilament proteins.**
Box plots showing the signals for the three recombinant neurofilament proteins (NfL, NfM, and NfH), produced by antibodies in indirect and sandwich ELISAs. The 13-0400 and 18934-1-AP are used as positive controls for NfL and NfH recombinant proteins. Ab, antibody; NfH, neurofilament heavy chain; NfL, neurofilament light chain; NfM, neurofilament medium chain; Rec, recombinant protein.

### Supplementary material

#### Assay performance validation

Stability assessments were carried out for two CSF samples. The obtained data revealed that NfM concentrations exhibited less than 20% variation when stored for up to three days at either 4 °C or room temperature. Additionally, changes in CSF NfM concentrations remained within an acceptable range of ±20% after undergoing five freeze-thaw cycles (Figure S2A). Dilution-adjusted concentrations of two CSF samples in the parallelism test were plotted (Figure S2B), and a 1:2 dilution was chosen as the minimum required dilution.

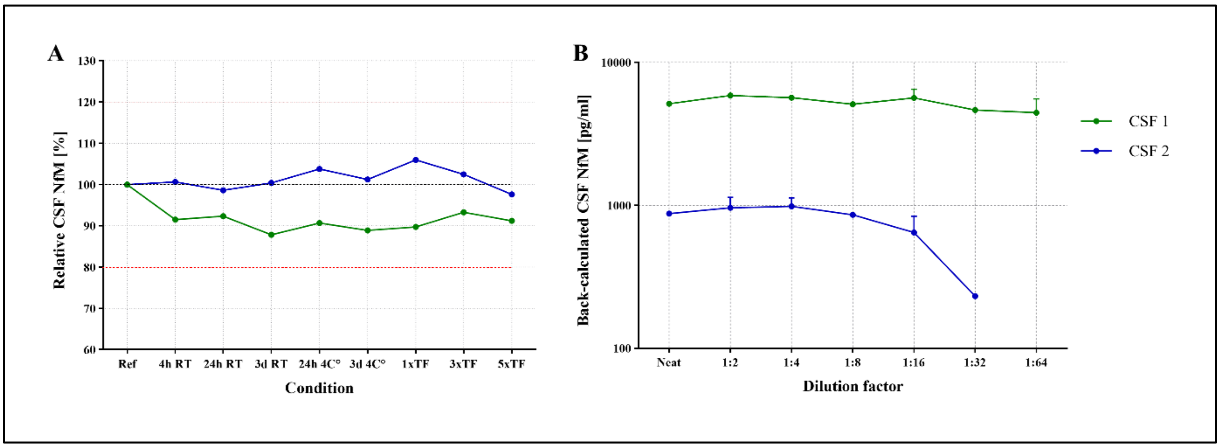


**Figure S2. NfM stability and parallelism assessment in CSF.**
(A) The stability of NfM in CSF was determined by comparing the relative content of NfM in two CSF samples after storage at room temperature or 4 °C and multiple freeze-thaw cycles, in comparison to the reference samples. Variations were found to be less than 20 %. CSF NfM remained stable after undergoing up to five freeze-thaw cycles and storage at room temperature or 4°C up to three days. (B) Back-calculated NfM concentrations within a serial dilution of two CSF samples. CSF, cerebrospinal fluid; NfM, neurofilament medium chain; TF, freeze-thaw cycle.

#### Neurofilaments ratio in diagnostic groups

The calculation of neurofilament ratios revealed significantly higher values for all three ratios in the AD (NfM/NfL: p<0.0001; NfM/NfH: p<0.0001; NfL/NfH: p=0.0283) and FTD (NfM/NfL: p<0.0001; NfM/NfH: p<0.0001; NfL/NfH: p<0.0001) cohorts compared to the CTRL group. In contrast, the ALS cohort showed significant differences from CTRL only for the NfM/NfL (p=0.0159) and NfM/NfH (p=0.0247) ratios. Additionally, the NfM/NfH (p=0.0008) and NfL/NfH (p<0.0001) ratios were significantly different in the ALS cohort compared to the CTRL.DD group (Figure S3A). Furthermore, pairwise comparisons within FTD subgroups revealed significant lower levels in lvPPA compared to nfvPPA (NfM/NfL: p=0.0274; NfM/NfH: p=0.0064; NfL/NfH: p=0.0169), and to svPPA (NfM/NfL: p=0.0152; NfM/NfH: p=0.0006; NfL/NfH: p=0.0006). Additionally, svPPA cohort showed significant higher values compared to bvFTD (NfM/NfH: p=0.0385; NfL/NfH: p=0.0049) and to nfvPPA (NfL/NfH: p=0.0041). No significant differences were observed within the LBD subgroups (Figure S3B).

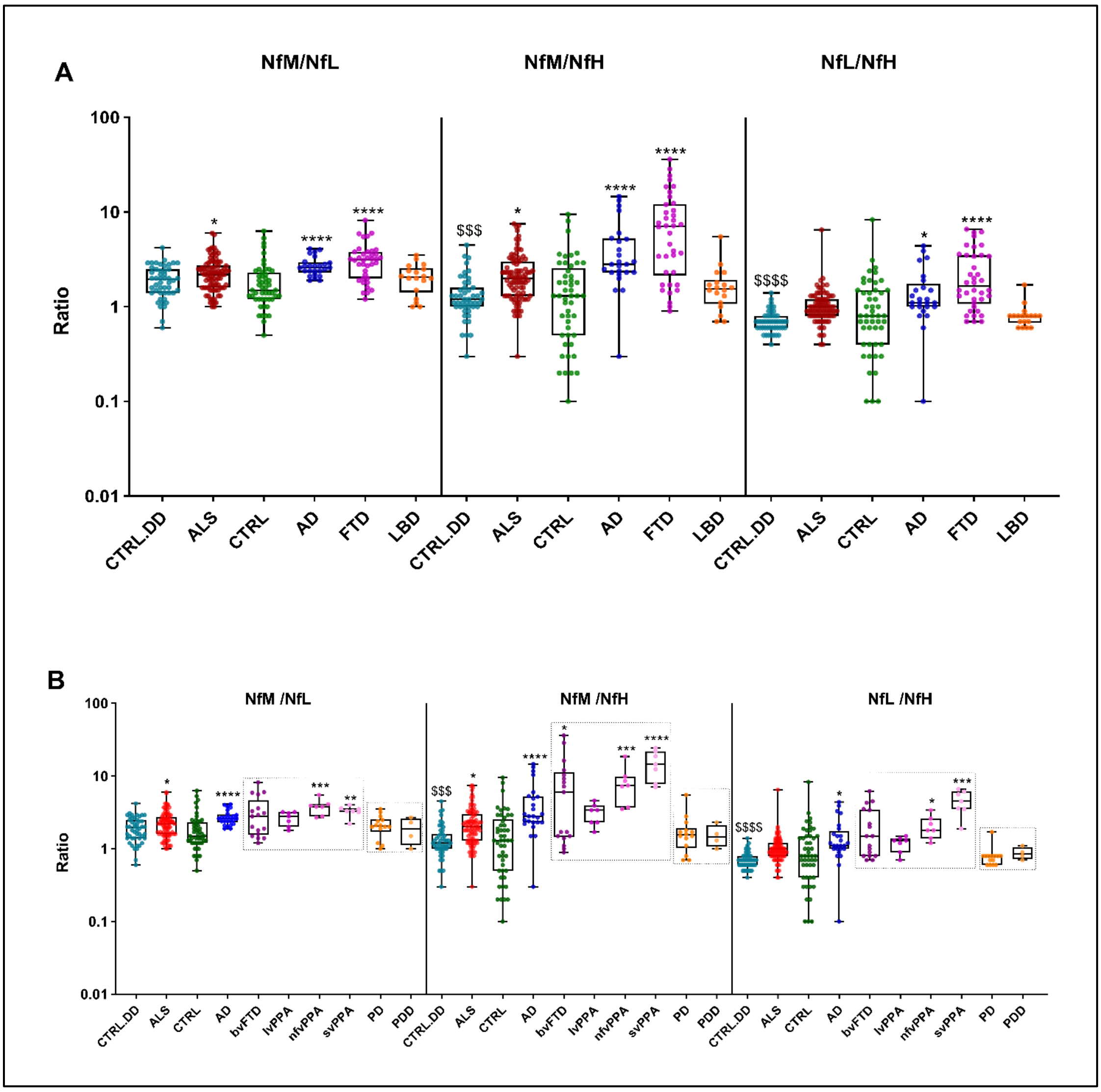


**Figure S3. Ratio of neurofilament subunits across different diagnostic groups.**

Ratios of CSF NfM, NfL and NfH concentrations in the diagnostic groups (A) and in the extended diagnostic groups (B). Statistically significant differences between the patient cohorts and CTRL are indicated with star symbols (*), while the comparison between ALS patients and the CTRL.DD cohort is marked with dollar signs ($). Additional significant differences within the patient cohorts are observed with the following p-values; NfM/NfL (ALS vs FTD: p=0.0092; FTD vs LBD: p=0.0359), NfM/NfH (ALS vs FTD: p= 0.0003; AD vs LBD: p=0.0093; FTD vs LBD: 0.0002; ALS vs nfvPPA: p=0.0452; ALS vs svPPA: p=0.0059; nfvPPA vs PD: p=0.0097; svPPA vs PD: 0.0015), and NfL/NfH (ALS vs FTD: p=0.0005; AD vs LBD: p=0.0079; FTD vs LBD: p<0.0001; ALS vs svPPA: p=0.0057; AD vs PD: p=0.0229; bvFTD vs PD: p=0.0273; nfvPPA vs PD: p=0.0032; svPPA vs PD: p<0.0001). Displayed are the median concentration, the 25% and 75% percentiles and whiskers from minimum to maximum. Groups were compared by Kruskal-Wallis test and Dunns post hoc test.( * p< 0.05; ** p< 0.01;

### Supplementary material

*** p< 0.001; **** p<0.0001; and $$$$ p<0.0001) AD, Alzheimer's disease; ALS, amyotrophic lateral sclerosis; bvFTD, behavioral variant frontotemporal dementia; CTRL, non-neurodegenerative controls; CTRL.DD, control patients with initial diagnostic suspicion of ALS but finally diagnosed with another condition; CSF, cerebrospinal fluid; FTD, frontotemporal dementia; LBD, lewy body dementia; lvPPA, logopenic variant primary progressive aphasia; NfH, neurofilament heavy chain; NfL, neurofilament light chain; NfM, neurofilament medium chain; nfvPPA, non-fluent variant primary progressive aphasia; PD, Parkinson's disease; PDD, Parkinson's disease dementia; svPPA, semantic variant primary progressive aphasia.

**Table S1. Diagnosis and neurofilament concentrations of the CTRL.DD group**

| Diagnosis | N | f/m | Age (year) | CSF NfM (pg/mL) | CSF NFL (pg/mL) | CSF NfH (pg/mL) |
|---|---|---|---|---|---|---|
| Benign fasciculations | 6 | 1/5 | 49 (43-57) | 698 (465-1747) | 598 (324-795) | 873(645-1181) |
| brachial plexus injury | 3 | 1/2 | 57 (45-60) | 796 (687-1019) | 441 (355-463) | 751 (602-793) |
| HSP | 2 | 1/1 | 59 (56-62) | 1174 (1049-1298) | 696 (653-739) | 874 (770-979) |
| IBM | 3 | 2/1 | 71 (70-77) | 4353 (1587-4577) | 1092 (867-1516) | 1503 (1362-1533) |
| Multifocal motor neuropathy | 2 | 0/2 | 66 (66-67) | 1078 (879-1277) | 677 (582-772) | 1211 (1205-1216) |
| Myopathy | 7 | 3/4 | 54 (40-65) | 1109 (677-2721) | 482 (463-1091) | 829 (708-1423) |
| PNP | 13 | 3/10 | 71 (53-75) | 1819 (687-3622) | 801 (525-1343) | 1285 (681-1986) |
| Bilateral L5 radiculitis | 1 | 0/1 | 66 | 342 | 578 | 1010 |
| Cervical and lumbar disk herniation | 1 | 0/1 | 52 | 555 | 506 | 531 |
| Cervical disk prolaps | 1 | 1/0 | 56 | 659 | 424 | 891 |
| Fibromyalgia | 1 | 1/0 | 59 | 1106 | 501 | 714 |
| Inclusion body myositis | 1 | 0/1 | 43 | 503 | 305 | 521 |
| Mononeuritis multiplex | 1 | 0/1 | 79 | 2066 | 868 | 1275 |
| MS | 1 | 0/1 | 50 | 875 | 329 | 814 |
| Myelitis | 1 | 0/1 | 50 | 810 | 587 | 719 |
| Myositis | 1 | 1/0 | 78 | 1781 | 1176 | 978 |
| post-polio syndrome | 1 | 0/1 | 63 | 1304 | 671 | 1003 |

### Supplementary material

| | | | | | | |
|---|---|---|---|---|---|---|
| SMA | 1 | 0/1 | 54 | 856 | 433 | 639 |
| Somatic symptom disorder | 1 | 1/0 | 38 | 165 | 231 | 335 |

Data is reported as median (Interquartile range).

CTRL.DD, control patients with initial diagnostic suspicion of ALS but final diagnosis of different condition; CSF, cerebrospinal fluid; HSP, hereditary spastic paraplegia; IBM, Inclusion body myopathy; MS, multiple sclerosis; NfH, neurofilament heavy chain; NfL, neurofilament light chain; NfM, neurofilament medium chain; PNP, polyneuropathy; SMA, spinal muscular atrophy.
